# COVID-19: Can early home treatment with Azithromycin alone or with Zinc help prevent hospitalisation, death, and long-COVID-19? A review

**DOI:** 10.1101/2020.12.29.20248975

**Authors:** Philippe Lepere, Bruno Escarguel, Selda Yolartiran, Claude Escarguel

**Affiliations:** University of Geneva, Institute of Global Health, 24 rue du General-Dufour 1205 Geneva, Switzerland; Saint Joseph Hospital, COVID Unit, 26 Bd de Louvain 13008 Marseille, France; Association Biologie et Cooperation, 135 Traverse du Gaou 83140 Six-Fours, France

**Keywords:** COVID-19, azithromycin, early treatment, family practitioners

## Abstract

**Introduction:** The effects of the SARS-CoV-2 pandemic continues to disrupt health systems worldwide, leading to population lockdowns in many countries. Preventing hospitalisation, death and long-COVID-19 with repurposed drugs remains a priority. Hydroxychloroquine (HCQ) and azithromycin (AZM) are the most commonly used in ambulatory care, with divergent results. With the aim of decentralizing early treatment to family practitioners, we addressed the question: *Can early home treatment with AZM alone or with zinc help prevent hospitalisation, death, and long-COVID-19?*

**Methodology:** We conducted a scoping review of articles published from 31^st^ December 2019 to 5^th^ November 2020 in Pubmed, Google Scholar, MedRxiv, and BioRxiv databases, and a review of undergoing clinical trials published in the Clinicaltrial.gov database.

**Results:** Many studies report on outpatient treatment with a combination of AZM + HCQ versus AZM alone, and few studies propose the addition of Zinc (Zn) to AZM. In addition, we identified 5 clinical trials currently recruiting individuals for early outpatient treatment with AZM. However, we failed in identifying any study or clinical trial conducted with family practitioners responding to our question.

**Discussion:** The antiviral, anti-inflammatory, immunomodulatory benefits of AZM + Zn make this drugs combination a good candidate therapy to treat flu-like-COVID-19 and atypical pneumoniae. The antibacterial action of AZM can also help disrupting the bacteria/virus cooperation that is poorly documented. Considering pros and cons of macrolide use (including antimicrobial resistance), we call for early use of this therapy by family practitioners for home treatment of individuals presenting mild or moderate symptoms under rigorous scientific guidance to prevent hospitalisation, death and long-COVID.

## Introduction

The first case of coronavirus disease [1], a disease caused by the novel severe acute respiratory syndrome coronavirus 2 (SARS-CoV-2), was reported in the province of Wuhan, China, on 31^st^ December 2020. As of 5^th^ November 2020, over 48 million confirmed cases and 1,2 million deaths were reported by the World Health Organisation [2], leading to major disruptions in health systems and public lockdowns worldwide. In the absence of effective treatment, only reinforced prevention and early treatment of coinfections with repurposed drugs can disrupt viral transmission. Azithromycin [3], a macrolide antibiotic with antiviral, anti-inflammatory, and antibacterial properties seems to be a good treatment option. We reviewed studies published from 31^st^ December 2019 to 5^th^ November 2020 addressing the question: *Can early home treatment with AZM alone or with zinc help prevent hospitalisation, death, and long-COVID-19?*

### 5 steps of COVID-19 disease progression

The genome of the novel SARS-CoV-2 has been sequenced to study its host adaptation, viral evolution, infectivity, transmissibility, and pathogenicity [4]. For the purpose of our study, we have drown on the classification system proposed by Siddiqi et al. [5] to characterise the infection cycle as follows: Stage 1 – person-to-person infection via respiratory droplets produced when an infected individual coughs or sneezes (airborne transmission); stage 2 – viral penetration to host cells via two receptors: angiotensin-converting enzyme 2 (ACE2) and CD147 [6]; stage 3 – SARS-CoV-2 inhibits and evades the innate immune response and drives pathogenesis (viral replication provokes localised inflammation in the lung, leading to viral pneumonia; there is no clear boundary between the viral and inflammatory stages and they may overlap. At this stage, most patients need to be hospitalised for close observation and treatment); stage 4 – progression to acute respiratory stress associated with increased production of proinflammatory cytokines; stage 5 – the patient must be admitted to the intensive care unit as he/she is most likely to develop a cytokine storm and autoimmune disorders that can lead to death.

### Duration of infectiousness and symptoms

The duration of infection from symptom onset to recovery is approximately 10 days in non-severe cases [7]. Viral peak appears in the upper respiratory tract within the first week of symptom onset, and later in the lower respiratory tract in both asymptomatic and symptomatic infected individuals. Viral load clearance is faster in asymptomatic than in symptomatic patients. Generally, individuals recover within 3 weeks. However, the post-recovery course of the disease, including its physical and psychological sequelae, presents many unknowns. Around 10% of the patients who tested positive for the SARS-CoV-2 virus remained unwell for more than 3 weeks, and a small proportion did so for months [8]. Prolonged COVID-19 can induce long-term pulmonary disorders and have adverse effects on the heart, kidneys, digestive tract, or neural system. Age, comorbidities, history of smoking, length of hospitalisation, severity of the acute disease (such as the need for ICU admission), and the type of medications administered (such as antiviral or corticosteroid therapy) are the most important determinants of disease progression [9]. In addition, the consequences on mental health are underestimated [10].

### Preventive strategies and therapy

At stage 1, only preventive measures, such as washing hands, covering the mouth and nose when coughing and sneezing, refraining from shaking hands, wearing a mask, and physical distancing [11], have proven to be effective in partially controlling the spread of the epidemic by disrupting person-to-person transmission. Further prevention methods include testing, contact tracing of index cases, and isolation of positive cases [12, 13].

At stage 2, there is a need to develop drugs that can potentially block the host cell receptors ACE2 and CD147. AZM presents this capacity [6]. The combination of (hydroxychloroquine) HCQ + AZM has a synergic inhibitory effect on the replication of SARS-CoV1 and SARS-CoV2, which can be beneficial in the early stage of COVID-19 infection by reducing the viral load *in vitro* [14]. AZM occupies the ganglioside-binding domain of the spike protein and neutralises virus binding to lipid rafts, while HCQ covers the ganglioside surface and prevents virus-membrane interaction through a complementary mechanism [15].

At stage 3, efforts focused on blocking viral replication with antiretroviral drugs such as those against human immunodeficiency virus (HIV) or Ebola virus (e.g. remdesivir). Most commonly tested drugs were inhibitors of RNA polymerase, such as lopinavir, ritonavir, or darunavir targeting the transcription of the viral genome, and inhibitors of regulatory proteins such as remdesivir, ribavirin, or favipiravir, which target the translation of viral proteins. To date, only remdesivir has shown limited efficacy in reducing the length of stay in ICU from 15 to 11 days. However, on 19^th^ November 2020, due to the high number of reported adverse effects and its high cost, the WHO stated that remdesivir should not be used to treat hospitalised patients with COVID-19, regardless of disease severity [16]. In parallel, whether corticoid therapy should be initiated to stop the inflammatory process that could be concomitant with viral replication is still unclear. Since steroids are considered corrective, anti-inflammatory interventions to be administered later in the diseases course, the WHO[17] expressed its concern about early steroid use since there is little or no evidence of its effectiveness in COVID-19 patients.

At stage 4, it is important to the limit the production of cytokines, particularly interleukin 6 (IL6), and of interferons. Anti-inflammatory drugs such as IL6 inhibitors, corticosteroids, or tocilizumab may remediate severe damage and prevent a cytokine storm.

### Why is azithromycin a good candidate therapy?

AZM, a macrolide antibiotic, has a well-known safety profile; it is easily produced at a low cost as a generic drug and declared an essential medicine by the WHO [18]. It is distributed worldwide, making it compliant with the WHO’s policy for drug repositioning [19]. AZM is effective against gram-positive bacteria, some gram-negative bacteria, and many atypical bacteria. Common side effects include nausea, vomiting, diarrhoea, and upset stomach. Allergic reaction, such as anaphylaxis, QT prolongation, or diarrhoea caused by *Clostridium difficile* is possible. In the search for a safe and effective treatment of early mild or moderate COVID-19, AZM seems to be the most promising option.

AZM has antiviral, immunomodulatory, and clinical effects in the treatment of COVID-19 [20, 21]. At stage 2, AZM can occupy the ganglioside-binding domain of the spike protein and neutralise virus binding to lipid rafts. It also interferes with the ligand CD147 receptor interaction (antiviral action). At stages 4 and 5, AZM can reduce the synthesis of proinflammatory cytokines, and as an outcome, reduce the length of stay or the need for respiratory support during hospitalisation (immunomodulatory effect).

Finally, there is a paucity of literature on coinfection with bacterial species in COVID-19 patients. The most frequently isolated species are, in descending order, *Mycoplasma pneumoniae, Staphylococcus aureus, Legionella pneumophila, Haemophilus spp., Klebsiella spp., Pseudomonas aeruginosa, Chlamydia spp., S. pneumoniae, and Acinetobacter baumannii* [22]. The respiratory symptoms of patients with COVID-19 pneumonia admitted to the hospital with fever and dry cough can mimic those of atypical bacterial pneumonia, making it difficult to distinguish COVID-19 pneumonia from hospital-acquired and ventilator-associated pneumonia. Antibiotic treatment should be designed considering the possible side effects (e.g. QT prolongation, diarrhoea), local epidemiology of drug resistance, and impact of drug resistance on the patient [22]. Macrolide antibiotics, particularly AZM, remain an interesting option in specific conditions.

### Why consider treatment with Zinc?

Zinc (Zn) is well tolerated, known for its antioxidant, anti-inflammatory, immunomodulatory, and antiviral activities. Elderly people have an increased probability of zinc deficiency. In the elderly, low Zn status (serum Zn values <0.7 mg/L) represents a risk factor for pneumonia [23]. Inadequate Zn supply may predispose individuals to infectious diseases of the upper and lower respiratory tract. Although the therapeutic effects of Zn are inconsistent, evidence-based data indicate the efficiency of Zn supplementation in preventing pneumonia and its complications due to the anti-inflammatory properties of zinc [24, 25]. Recently, in vitro results indicated that low zinc levels favor viral expansion in SARS-CoV2 infected cells [26].

### Why promote ambulatory care?

With the continuous expansion of the pandemic and the resurgence of a second wave in Europe, health systems are facing many disruptions worldwide. In many countries, this situation has led to public lockdowns. To decongest hospitals, we think it is crucial to support frontline health workers and family practitioners in playing key roles in triage, early detection, and early treatment of patients with mild and moderate symptoms. Only the most vulnerable patients at risk of severe disease progression should be referred to the hospital.

### Methodology: the review

We adopted Arksey and O’Malley’s [27] five-stage framework for a scoping review: identifying the research question, identifying relevant results, selecting studies, charting data, and reporting results. We defined the following research question: *Can early home treatment with AZM alone or with zinc help prevent hospitalisation, death, and long-COVID-19?*

The scientific literature review was conducted by searching the online databases of PubMed and Google scholar using the following search terms and Booleans: (COVID-19 OR SARS-CoV-2 OR coronavirus) AND (azithromycin OR Zithromax) AND (outpatient OR ambulatory OR early treatment). Preprints were selected from bioRxiv, and medRxiv platforms. We accessed grey literature using the Google search engine. Materials published from 31^st^ December 2019 to 5^th^ November 2020 were searched. One researcher (PL) independently searched the databases and conveyed his findings to the co-authors. We also performed an advanced search on the Clinicaltrials.gov database using the keywords ‘COVID, SARS-CoV-2, azithromycin, and Zithromax’.

## Findings

### 1. Articles

A total of 350 articles were identified, of which 19 (17 peer-reviewed articles + 2 preprints) were selected for full review (see Fig. 1). The inclusion criteria were outpatient or ambulatory treatment, home treatment prescribed by general practitioners, use of AZM alone or with Zn, and duration of AZM treatment ≥ for 5 days. We failed to identify any study fulfilling all these criteria, from both peer-reviewed literature and preprints because none of these studies involved family practitioners. Most articles focused on *in vitro* action of AZM, review of opinions, and properties of AZM. We paid particular attention to two articles presenting a research protocol [28, 29] and one article in which the author [30] recommended formal clinical trials after having successfully treated more than 50 patients presenting flu-like symptoms, with AZM (500 mg at day 1 + 250 mg for the remaining 5 days). Clinical improvement was observed in all patients 24–48 hours after treatment initiation.

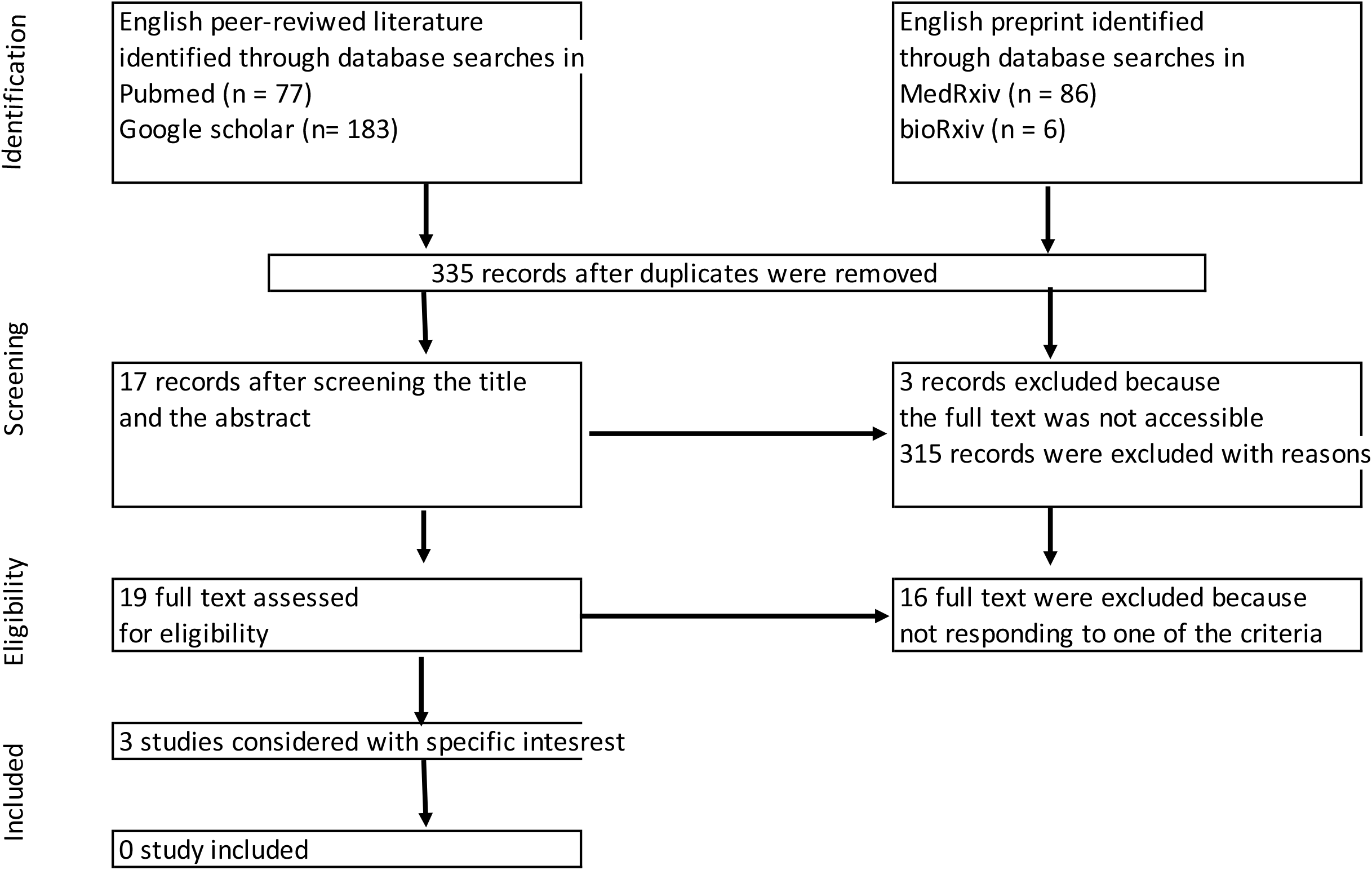

Many excluded articles (peer-reviewed studies, preprints, commentaries, letters, editorials, etc.) reported the effectiveness of a combination of HCQ + AZM administered to patients at different stages of the disease, from outpatient treatment to ICU care. Gautret et al. [31] were the first to highlight the effectiveness of dual therapy, showing better results with HCQ + AZM than with HCQ alone, under rigorous cardiological surveillance, in reducing hospitalisation and mortality. Many other studies concluded that given the efficacy of HCQ + AZM in early outpatient treatment, the evidence on the use of HCQ alone or HCQ + AZM in inpatients is irrelevant to its use in high-risk outpatients in early stages of the disease [32, 33]. In addition, triple therapy with HCQ + AZM + Zn improved the outcomes and reduced the duration of hospitalisation [34]. However, most of these studies had a small sample size, non-robust methodology, different measures of the primary endpoint, and inconclusive results.

We found several press releases on Google showing encouraging results [35, 36] with the use of AZM, but we did not consider those press releases as evidence because of methodological limitations.

### 2. Protocols for clinical trials

We identified a total of 3,904 NIH registered clinical trials on the ClinicalTrials.gov database for COVID-19. Through an advanced search, we selected 121 studies, out of which 88 were on going. We identified 10 clinical trials on the effect of AZM, with at least one arm comprising patients treated with AZM alone for a minimum duration of 5 days, and one clinical trial proposing a single dose of AZM. Five trials were hospital-based and six focused on ambulatory care. We selected the latter six trials (see Table 1).

**Table 1.** identified clinical trial having at least one arm with AZM and recruiting infected individuals at outpatient units.

| Principal investigator | Study | Status | Design | # patients | Remarks |
| --- | --- | --- | --- | --- | --- |
| Alaa Rashad<br>South-Vally<br>university, Egypt | Clarithromycin versus<br>azithromycin in treatment of<br>mild COVID-19 infection | Completed<br>(May to July<br>2020)<br>No results<br>available | 3 arms double-blinded randomized<br>control trial | 300 participants in total<br>107 participants aged<br>45.9 ± 18 in AZM arm | AZM 500 mg/24h for 7<br>days<br>Primary outcome: time to<br>complete resolution of<br>fever |
| Paul Little<br>University of<br>Oxford, UK | ATOMIC2: a Multi-centre<br>open-label two-arm<br>randomised superiority<br>clinical trial of azithromycin<br>versus usual care in<br>ambulatory COVID-19 | Recruiting<br>No results<br>available | Open label, two-arms, randomized<br>superiority trial | Adults (aged 18+)<br>presenting to hospital<br>with clinically-<br>diagnosed COVID-19<br>infection managed<br>initially as outpatient<br>800 participants<br>Currently in phase III | Daily dose of 500mg for<br>14 days<br>Primary outcomes<br>measured at Day 28:<br>proportion progressing to<br>respiratory failure |
| Thomas M.<br>Lietman<br>University of<br>California, USA | ACTION: Azithromycin for<br>COVID-19 treatment in<br>outpatients nationwide. | Recruiting<br>No results<br>available | Individually randomized telemedicine-<br>based, placebo-controlled trial | 2271 participants<br>In progress | Primary outcome:<br>efficacy of a single 1.2 g<br>dose of ATM for<br>prevention of disease<br>progression to<br>hospitalisation |
| Brandon Web<br>University of Utah,<br>USA | HyAzOUT:<br>Hydroxychloroquine vs.<br>azithromycin for outpatients<br>in Utah: a prospective<br>programmatic trial | Recruiting<br>No results<br>available | Randomized clinical trial open label with<br>two arms | Adults aged > 44 years<br>1550 patients<br>Recruitment in process | ATM 500 mg PO on day<br>1 plus 250 mg on days 2-5<br>Primary outcome:<br>admitted to a hospital<br>within 14 days of<br>enrolment |
| Jean-Philippe<br>Lanoix,<br>Centre hospitalier<br>universitaire<br>d'Amiens, France | AMBU-COVID : Proactive<br>care of ambulatory COVID-19<br>patients | No results<br>available | Randomised clinical trial open label | 64 participants<br>In progress | ATM 500mg on day 1,<br>then 250 mg the<br>following 4 days PO<br>Primary outcome: length<br>of symptom duration with<br>ATM treatment |
| Shenoor Azhar<br>University of health<br>science Lahore,<br>Pakistan | PROTECT: Pakistan<br>Randomized and<br>Observational Trial to<br>Evaluate Coronavirus<br>Treatment of<br>Hydroxychloroquine,<br>Oseltamivir and Azithromycin<br>to treat newly diagnosed<br>patients with COVID-19<br>infection who have no<br>comorbidities like diabetes<br>mellitus | Recruiting<br>No results<br>available | Double-blinded randomized trial.<br>Seven comparator groups: Each drug<br>(Hydroxychloroquine Phosphate/Sulfate,<br>Oseltamivir and Azithromycin) given as<br>monotherapy (three groups);<br>combinations of each of two drugs (three<br>groups); and a final group on triple drug<br>regimen. | 500 participants<br>Newly diagnosed 18+<br>patients, either<br>hospitalized or in self-<br>isolation<br>Number of participants<br>not determined | Azithromycin (500 mg<br>orally daily on day 1,<br>followed by 250 mg<br>orally twice a day on<br>days 2-5) alone<br>Primary outcome: turning<br>PCR test to negative on 7<br>days follow-up |

#### ACTION

This is the most innovative clinical trial on home self-treatment using mobile phones for follow-up. However, in the cohort study, AZM was administered as a single dose. This trial did not study the acceptability and adherence to a 6-day home self-treatment regime (AZM 500 mg on day 1 + 250 mg for the 5 following days).

#### ATOMIC2

The AZM dose regimen and duration of treatment were similar to those recommended in the UK for Lyme disease (500 mg daily for 14 days).

#### HyAzOUT

This study involved individuals aged 44+ years who presented to the hospital. This study will compare the benefits of HCQ and AZM.

#### AMBU-COVID

The Clinicaltrial.gov website has not been updated since the announcement of this trial, and we did not find further information on PubMed, Google Scholar, medRxiv and bioRxiv databases. Further, the sample size may not allow us to conclusively answer the research question.

#### PROTECT

This clinical trial provided relevant information from low- and middle-income countries. However, the primary endpoint was negative PCR results at day 7, which may not be the most relevant endpoint, as HCQ and AZM may delay virus clearance to more than 28 days after symptom onset [37]. This delay in viral clearance is correlated with older age.

Clarithromycin versus azithromycin in treatment of mild COVID-19 infection study completed. Low size of AZM arm, AZM dose regime (500 mg) duration of treatment (7 days) and primary endpoint (time to complete resolution of fever) will not allow to respond to our research question.

These trials proposed different AZM dose regimens, from a single dose to 10 days of treatment. If one or more trials provide evidence of AZM’s efficacy in a particular population or setting, further studies will be needed to provide conclusive data on population, settings, and dose regimens (28). Furthermore, none of these trials involved family practitioners. Hence, we did not find information on the added value of decentralising early detection and care to family practitioners, who best know their patient’s habits, behaviours, comorbidities, etc. Further, the trials had different primary outcomes. Hence, the results cannot be compared. Finally, to the best of our knowledge, these trials did not provide information on the prevalence of coinfections before initiating the treatment or after recovery.

## Discussion

We outlined five stages of COVID-19 disease progression. COVID-19 response strategies should consider these stages and propose appropriate strategies such as testing, contact tracing, isolation of confirmed cases with home-based early treatment, hospitalisation of severe cases, and immunisation. In this discussion, we will focus on early home treatment that could significantly reduce the number of infected people needing hospital-based care.

He Z. et al. [38] analysed 2,034 COVID-19 studies registered in ClinicalTrials.gov as of 18th June 2020 and reported that the five most frequently tested drugs were HCQ (n = 148), AZM (n = 46), tocilizumab (n = 29), lopinavir (n = 20), and ritonavir (n =20). We did not embark on such a deep review, but from the 3,904 registered trials, as of 5^th^ November 2020, a quick scan showed that HCQ and AZM are still highly preferred, with 262 records mentioning the use of HCQ alone or in combination with AZM. Eleven studies proposed the use of AZM alone: six focused on the hospital inpatient level, and five, on the outpatient level. Despite being a promising medication, there is a paucity of data on the use of AZM alone in treating COVID-19. In addition, we failed to find any research protocol involving family practitioners.

Scientific knowledge on COVID-19 has been increasing rapidly, but as of 20^th^ November 2020, there is still no antiviral with proven efficacy; hence, the world is placing it hope on a vaccine. Press releases announced the start of trials on promising vaccines produced by Pfizer/BioNTech [39], Moderna, Spoutnick, and Sinovac. However, considering the uncertainties about long-term efficacy, the level of protection by age (e.g. will older people with a deficient immune response be sufficiently protected?), reactogenicity and side effects of mRNA vaccines, and logistic conditions (cold storage, production, etc.), it is too early to consider these vaccines as the ‘magic bullet’. Both vaccines and early treatment are complementary and necessary tools in a comprehensive package of prevention aimed at preventing hospitalisation, death, and long-COVID. In Europe, and particularly in France, the response strategy for the COVID-19 epidemic failed to involve frontline health workers. A paradigm change is urgently needed to shift from a 100% hospital-based approach to a family practitioner-based strategy to avoid further disruption of the health system and lockdowns. In addition, recent data suggests focusing on those with long-COVID-19, also termed as COVID long-haulers [40]. The long-term physical and mental consequences of the disease are still unknown.

Dry nose, loss of taste and/or smell, and muscle pain are frequent complaints of COVID-19 patients who visit their family practitioner [41, 42], while hospitalised patients [4] usually complain of fever, dry cough, dyspnoea, chest pain, fatigue, and myalgia. Almost 95% of the infected people present mild or moderate symptoms that do not necessitate hospital-based care. Approximately 5% of the patients with COVID-19 and 20% of those hospitalised experience severe symptoms necessitating intensive care [43]. Therefore, family practitioners should play a central role in triage, early treatment of patients with mild and moderate symptoms, and referral to the hospital when early treatment fails or for most-at-risk vulnerable individuals. However, this approach presents two major challenges: the need for a drug with proven effectiveness and early detection of COVID-19 symptoms for early care. As mentioned, HCQ + AZM is the most common drug therapy used for early treatment. However, when decentralising early care to family practitioners, it might not be opportune to consider HCQ because i) even if its efficacy is still controversial, it seems to work only when administered early enough [44], and ii) it can have adverse effects requiring close monitoring. Clinical trials of HCQ or HCQ + AZM arms usually enrolled patients and initiated treatment at very early stages of infection, while patients in more advanced stages of COVID-19 with mild or moderate symptoms usually visit their general practitioners. At this particular stage, the use of HCQ could be counterproductive given its immunomodulating effect. When prescribed at an advanced stage of the disease, dual therapy has little effect; at this stage, only strong anti-inflammatory and/or anticoagulant treatments can help the patients [45]. In addition, in these late stages, the HCQ + AZM combination can be toxic in patients whose cardiac status is compromised [46]. HCQ appears to be the main driver of cardiac toxicity and not AZM itself, which is consistent with evidence that macrolides are not associated with an increased risk of cardiac events [20].

For these reasons, it is appropriate to focus on AZM, which is considered to be effective if administered less than 7 days after viral infection; it showed no adverse effects that cannot be monitored at the family practitioner level. In France, several groups of family practitioners, known as the *‘Laissons les médecins prescrire’ and ‘Azithro d’hospitalisation’* Alliances, claim to have obtained encouraging results with a small number of COVID-19 patients referred to the hospital after AZM treatment. These Alliances recommend the involvement of family practitioners and using AZM under rigorous scientific guidance. To support these groups of doctors, we (BE) promote the therapeutic algorithm (see Fig. 2) based on their own experience [47] and findings from the literature [20, 21, 48-51]. In parallel, the Association for the Prevention and the Management of Sanitary Crisis (UGPS) that represents individuals who have recovered from COVID-19 or are affected by it, supports these Alliances and is advocating for clinical trials. However, even if randomized control trials are considered as the ‘gold standard’, the body of arguments in favour of early treatment with AZM makes an RCT ethically questionable [52]. Considering that a well-designed cohort study can provide powerful results, we suggest mobilising the physicians of the above-mentioned Alliances for an observational study that could allow to address the research question in examining the early home treatment outcomes very quickly and easily.

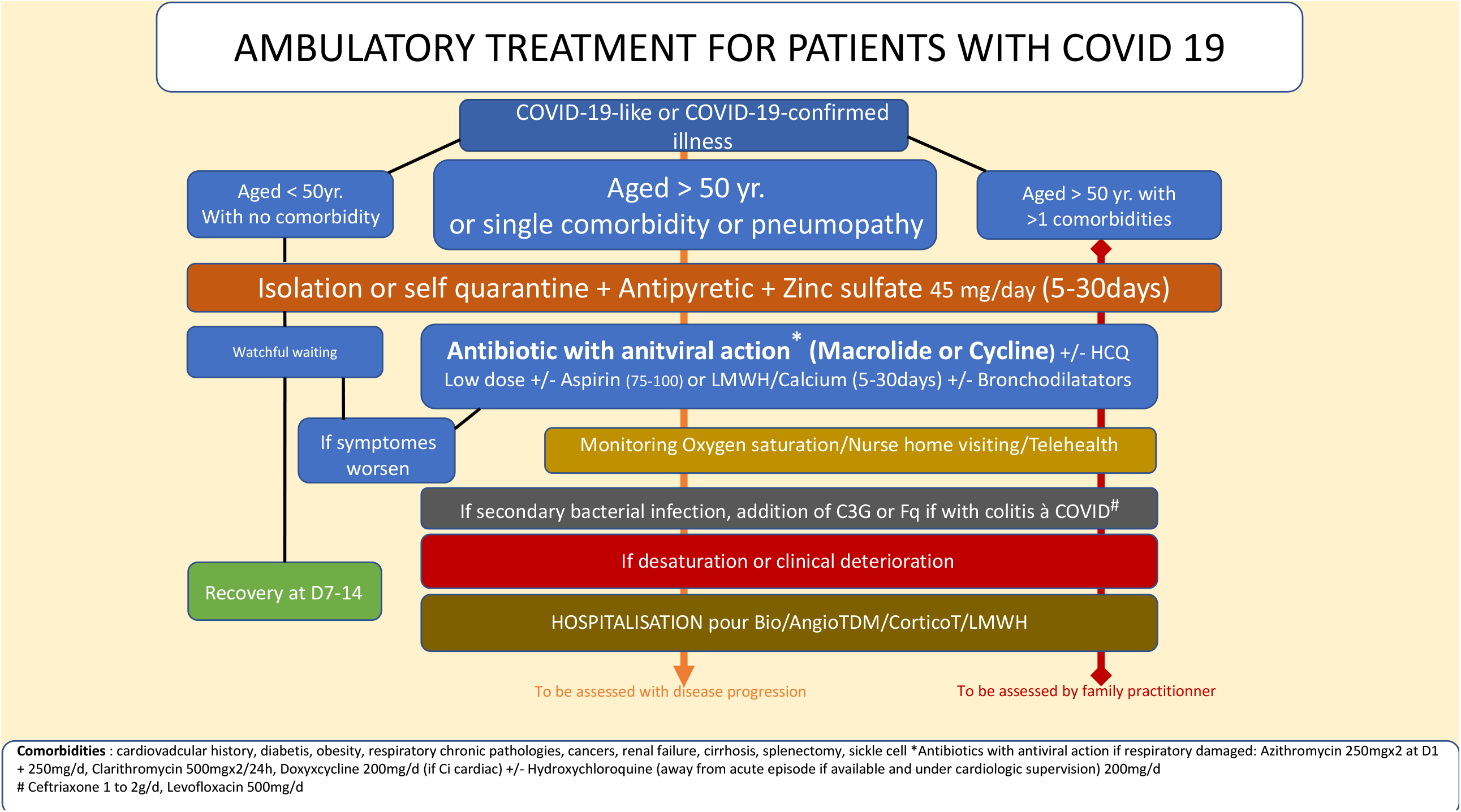

We need to be aware of new data that can be applied to the early ambulatory treatment of COVID-19. The role of *Mycoplasma pneumoniae* (MP) remains unclear. Lansbury et al. [53] do not recommend the routine use of antibiotics in the management of confirmed COVID-19 infection because the level of MP–SARS-CoV-2 coinfection is too low, while Nicolson and de Mattos [54] suggested that the severity of signs and symptoms in progressive COVID-19 patients could be partly due to MP and other bacteria. We recommend considering the role of MP–SARS-CoV-2 coinfection and the need to address it to prevent severe disease progression [55]. As demonstrated in a previous work (by CE, who obtained the patent EP0349473b1), certain mycoplasmas contribute to the explosive amplification of the replication of certain RNA viruses, such as the respiratory syncytial virus (RSV) *in vitro*. Bacteria such as *Chlamydia pneumoniae (CP), MP, Borrelia burgdorferi (BB), and Legionella pneumophila (LP)* are generally present in the pulmonary microbiota, hidden intracellularly in a quiescent state [56]. They participate in the development of local immune disorders leading to superinfection. Studies on the prevalence of *MP, CP, BB*, and *LP* present contradictory findings, considering the difficulty in isolating *MP*. When studying the seasonal distribution of *MP* among patients presenting with a mild acute respiratory symptom caused by the influenza virus A or B, or RSV, Layani-Milon et al. showed [57] the co-circulation of MP and viral strains of influenza A or B virus or RSV (see Fig. 3). The authors noted that every year, at least one peak of MP infection is observed in late autumn (October to December), and it varies in duration and intensity. Therefore, *MP* can superinfect patients presenting with viral infections, either in the early stage of infection (the first 3 days) or in the late stage during the recovery of respiratory cells. To conclude, this hypothesis favours early treatment of COVID-19 patients with AZM.

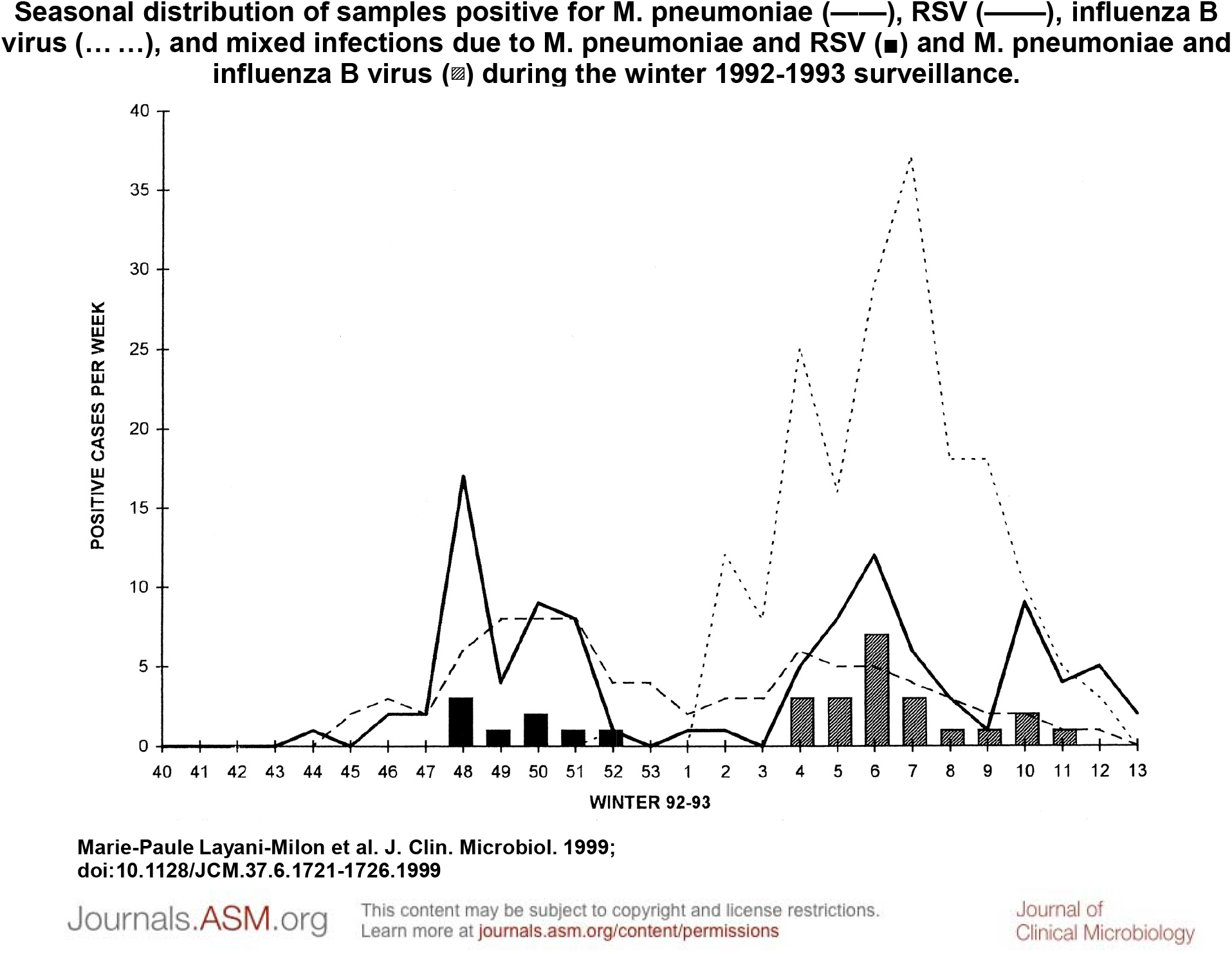

AZM is known to induce antimicrobial resistance at variable levels in different countries and regions [58], and antimicrobial resistance (AMR) is a matter of major concern [59]. Use of AZM and other antibiotics in the management of COVID-19 should be balanced against the risk of AMR. The paucity of available data makes it difficult to predict the impact that this pandemic may have on antimicrobial stewardship programmes and long-term rates of AMR [60]. The deaths from COVID-19 may overtake the deaths from AMR for 2020, but the estimated annual number of deaths from AMR of 10 million by 2050 may be higher than the death toll from the entire COVID-19 pandemic [61]. We encourage adopting a positive approach and considering the impacts of COVID-19 and AMR. For instance, behavioural interventions, including physical barriers, to prevent the spread of SARS-CoV-2, will likely decrease the spread of other infections and the use of antimicrobials [61]. Such interventions focusing on hand hygiene may have a considerable impact on AMR if adopted in the long term by individuals in their daily life and health workers in health facilities worldwide. Family practitioners can promote this. In addition, the high number of hospitalised SARS-CoV-2 infected individuals colonised with *carbapenemase-producing Enterobacteriaceae (CPE)/VRE/MRSA/Clostridioides difficile* increases the risk of nosocomial transmission within the hospitals [60]. Early home treatment with AZM will inevitably reduce the risk of hospital transmission and future consumption of antibiotics. This risk assessment should be addressed in collaboration with veterinary doctors and patient associations, as AMR is also caused by overuse of antibiotics in agriculture, animal, and human health. Key monitoring indicators and information to be collected are to be rapidly defined. Considering the interrelated emergencies resulting from COVID-19 and AMR, it could be opportune to involve veterinary doctors and representatives of patient associations in the national task forces that monitor the COVID-19 response at the country level.

## Conclusion

Our scoping review did not reveal evidence of early home treatment with AZM alone or associated with zinc be a reliable option to prevent hospitalisation, death, and long-COVID-19. We failed to identify studies involving family practitioners as frontline healthcare workers administering AZM treatment.

With the second wave of the COVID-19 pandemic in Europe, we suggest shifting from a hospital-based approach to a family practitioner-based approach with early home treatment that prevents hospitalisation, death, and long-term care. AZM shows the potential to respond to this objective. Research institutions should provide the necessary support to family practitioners in providing early home treatment with AZM +/-Zn under rigorous guidance. We suggest that national task forces managing COVID-19 response at the country level consider the insights of family practitioners, associations of COVID-19 patients who have recovered, and veterinary doctors.

## Data Availability

This article is a scoping review and all data are accessible on internet

## Bibliography

1. Recovery - Randomised Evaluation of COVID-19 Therapy. No clinical benefit from use of hydroxychloroquine in hospitalised patients with COVID-19 2020 [Available from: https://www.recoverytrial.net/news/statement-from-the-chief-investigators-of-the-randomised-evaluation-of-covid-19-therapy-recovery-trial-on-hydroxychloroquine-5-june-2020-no-clinical-benefit-from-use-of-hydroxychloroquine-in-hospitalised-patients-with-covid-19.

2. The World Health Organization. WHO coronavirus disease (COVID-19) dashboard 2020 [Available from: https://covid19.who.int.

3. Stringhini SS, Wisniak AA, Piumatti GG, Azman ASA, Lauer SAS, Baysson HH, et al. Seroprevalence of anti-SARS-CoV-2 IgG antibodies in Geneva, Switzerland (SEROCoV-POP): a population-based study. Lancet (British edition). 2020.

4. Harapan HH, Itoh NN, Yufika AA, Winardi WW, Keam SS, Te HH, et al. Coronavirus disease 2019 (COVID-19): A literature review. Journal of infection and public health. 2020;13(5):667–73.

5. Siddiqi HKH, Mehra MRM. COVID-19 illness in native and immunosuppressed states: A clinical-therapeutic staging proposal.The Journal of heart and lung transplantation. 2020;39(5):405–7.

6. Ulrich HH, Pillat MM. CD147 as a Target for COVID-19 Treatment: Suggested Effects of Azithromycin and Stem Cell Engagement. Stem Cell Reviews and Reports. 2020;16(3):434–40.

7. Cevik M, Tate M, Lloyd O, Maraolo AE, Schafers J, Ho A. SARS-CoV-2, SARS-CoV, and MERS-CoV viral load dynamics, duration of viral shedding, and infectiousness: a systematic review and meta-analysis. The Lancet Microbe.

8. Hamming I, Timens W, Bulthuis M, Lely A, Navis Gv, van Goor HJTJoPAJotPSoGB, et al. Tissue distribution of ACE2 protein, the functional receptor for SARS coronavirus. A first step in understanding SARS pathogenesis. 2004;203(2):631–7.

9. Salehi SS, Reddy SS, Gholamrezanezhad AA. Long-term Pulmonary Consequences of Coronavirus Disease 2019 (COVID-19): What We Know and What to Expect. Journal of thoracic imaging. 2020;35(4):W87–W9.

10. Arora TT, Grey II. Health behaviour changes during COVID-19 and the potential consequences: A mini-review. Journal of Health Psychology. 2020;25(9):1155–63.

11. Chu DK, Akl EA, Duda S, Solo K, Yaacoub S, Schünemann HJ, et al. Physical distancing, face masks, and eye protection to prevent person-to-person transmission of SARS-CoV-2 and COVID-19: a systematic review and meta-analysis. The Lancet.

12. Steinbrook RR. Contact Tracing, Testing, and Control of COVID-19-Learning From Taiwan. JAMA Internal Medicine. 2020;180(9):1163–4.

13. He ZZ. What further should be done to control COVID-19 outbreaks in addition to cases isolation and contact tracing measures? BMC medicine. 2020;18(1):80-.

14. Andreani JJ, Le Bideau MM, Duflot II, Jardot PP, Rolland CC, Boxberger MM, et al. In vitro testing of combined hydroxychloroquine and azithromycin on SARS-CoV-2 shows synergistic effect. Microbial pathogenesis. 2020;145:104228-.

15. Fantini JJ, Chahinian HH, Yahi NN. Synergistic antiviral effect of hydroxychloroquine and azithromycin in combination against SARS-CoV-2: What molecular dynamics studies of virus-host interactions reveal. International journal of antimicrobial agents. 2020;56(2):106020-.

16. WHO guideline development group advises against use of remdesivir for COVID-19 [press release]. 20 November 2020 2020.

17. The World Health Organization. Corticosteroids for COVID-19. Geneva: The World Health Organization,; 2020 2 September 2020.

18. The World Health Organization. WHO model list of essential medicines - 21st list, 2019 2019 [Available from: https://www.who.int/publications/i/item/WHOMVPEMPIAU2019.06.

19. Singh TU, Parida S, Lingaraju MC, Kesavan M, Kumar D, Singh RK. Drug repurposing approach to fight COVID-19. Pharmacological reports: PR. 2020;72(6):1479–508.

20. EcheverrÌa Esnal DD, Martin Ontiyuelo CC, Navarrete Rouco MEM, De-Antonio CuscÛ MM, Ferr·ndez OO, Horcajada JPJ, et al. Azithromycin in the treatment of COVID-19: a review. Expert Review of Anti-infective Therapy. 2020:1–17.

21. Damle B, Vourvahis M, Wang E, Leaney J, Corrigan B. Clinical Pharmacology Perspectives on the Antiviral Activity of Azithromycin and Use in COVID-19. 2020;108(2):201–11.

22. Fattorini LL, Creti RR, Palma CC, Pantosti AA. Bacterial coinfections in COVID-19: an underestimated adversary. Annali dell’Istituto superiore di sanit‡. 2020;56(3):359–64.

23. Barnett JBJ, Hamer DHD, Meydani SNS. Low zinc status: a new risk factor for pneumonia in the elderly? Nutrition Reviews. 2010;68(1):30–7.

24. Skalny AVA, Rink LL, Ajsuvakova OPO, Aschner MM, Gritsenko VAV, Alekseenko SIS, et al. International Journal of Molecular Medicine. 2020;46(1):17–26.

25. Pal AA, Squitti RR, Picozza MM, Pawar AA, Rongioletti MM, Dutta AKA, et al. Zinc and COVID-19: Basis of Current Clinical Trials. Biological Trace Element Research. 2020.

26. Early Description of Coronavirus 2019 Disease in Kidney Transplant Recipients in New York. J Am Soc Nephrol. 2020;31(6):1150–6.

27. Arksey H, O’Malley L. Scoping studies: towards a methodological framework. International journal of social research methodology. 2005;8(1):19–32.

28. Hinks TSCTSC, Barber VSV, Black JJ, Dutton SJS, Jabeen MM, Melhorn JJ, et al. A multicentre open-label two-arm randomised superiority clinical trial of azithromycin versus usual care in ambulatory COVID-19: study protocol for the ATOMIC2 trial. Trials. 2020;21(1):718-.

29. Akram JJ, Azhar SS, Shahzad MM, Latif WW, Khan KSK. Pakistan Randomized and Observational Trial to Evaluate Coronavirus Treatment (PROTECT) of Hydroxychloroquine, Oseltamivir and Azithromycin to treat newly diagnosed patients with COVID-19 infection who have no comorbidities like diabetes mellitus: A structured summary of a study protocol for a randomized controlled trial. Trials. 2020;21(1):702-.

30. Schwartz RAR, Suskind RMR. Azithromycin and COVID-19: Prompt early use at first signs of this infection in adults and children, an approach worthy of consideration. Dermatologic Therapy. 2020;33(4):e13785–e.

31. Gautret PP, Lagier JJ-C, Parola PP, Hoang VTV, Meddeb LL, Mailhe MM, et al. Hydroxychloroquine and azithromycin as a treatment of COVID-19: results of an open-label non-randomized clinical trial. International journal of antimicrobial agents. 2020:105949-.

32. Risch HAH. Early Outpatient Treatment of Symptomatic, High-Risk COVID-19 Patients That Should Be Ramped Up Immediately as Key to the Pandemic Crisis. American journal of epidemiology. 2020;189(11):1218–26.

33. Brown SM, Peltan I, Kumar N, Leither L, Webb BJ, Starr N, et al. Hydroxychloroquine vs. Azithromycin for Hospitalized Patients with COVID-19 (HAHPS): Results of a Randomized, Active Comparator Trial. Annals of the American Thoracic Society. 2020.

34. Derwand R, Scholz M, Zelenko V. COVID-19 outpatients: early risk-stratified treatment with zinc plus low-dose hydroxychloroquine and azithromycin: a retrospective case series study. Int J Antimicrob Agents. 2020;56(6):106214.

35. Coronavirus. Trois médecins français satisfaits après avoir testé un nouveau traitement [press release]. 14 April 2020 2020.

36. Un médecin mosellan constate l’efficacité d’un protocole à base d’azithromycine [press release]. 11 April 2020 2020.

37. Saleemi SA, Alrajhi A, Alhajji M, Alfattani A, Albaiz F. Time to negative PCR from symptom onset in COVID-19 patients on Hydroxychloroquine and Azithromycin - A real world experience. 2020:2020.08.05.20151027.

38. He Z, Erdengasileng F, Luo X, Xing A, Charness N, Bian J. How the clinical research community responded to the COVID-19 pandemic: An analysis of the COVID-19 clinical studies in ClinicalTrials.gov. 2020:2020.09.16.20195552.

39. Polack FPF, Thomas FPSJ, Kitchin SJN, Absalon JNJ, Gurtman AJA, Lockhart SAS, et al. Safety and Efficacy of the BNT162b2 mRNA Covid-19 Vaccine. The New England Journal of Medicine. 2020.

40. Baig AMA. Chronic COVID Syndrome: Need for an appropriate medical terminology for Long-COVID and COVID Long-Haulers. Journal of Medical Virology. 2020.

41. Sebo P, Tudrej B, Lourdaux J, Cuzin C, Floquet M, Haller DM, et al. Clinical characteristics of SARS-CoV-2 patients: a French cross-sectional study in primary care. 2020.

42. Tudrej BB, Sebo PP, Lourdaux JJ, Cuzin CC, Floquet MM, Haller DMD, et al. Self-Reported Loss of Smell and Taste in SARS-CoV-2 Patients: Primary Care Data to Guide Future Early Detection Strategies. Journal of General Internal Medicine. 2020;35(8):2502–4.

43. Wiersinga WJ, Rhodes AA, Cheng ACA, Peacock SJS, Prescott HCH. Pathophysiology, Transmission, Diagnosis, and Treatment of Coronavirus Disease 2019 (COVID-19): A Review. JAMA: Journal of the American Medical Association. 2020;324(8):782–93.

44. Million MM, Lagier JJ-C, Gautret PP, Colson PP, Fournier PP-E, Amrane SS, et al. Early treatment of COVID-19 patients with hydroxychloroquine and azithromycin: A retrospective analysis of 1061 cases in Marseille, France. Travel Medicine and Infectious Disease. 2020:101738-.

45. Driggin EE, Madhavan MVM, Bikdeli BB, Chuich TT, Laracy JJ, Biondi Zoccai GG, et al. Cardiovascular Considerations for Patients, Health Care Workers, and Health Systems During the COVID-19 Pandemic. Journal of the American College of Cardiology. 2020;75(18):2352–71.

46. Magagnoli JJ, Narendran SS, Pereira FF, Cummings TT, Hardin JWJ, Sutton SSJ, et al. Outcomes of hydroxychloroquine usage in United States veterans hospitalized with Covid-19. medRxiv. 2020.

47. Christophe Brette, editor COVID-19: complications par pneumopathies. hypothèse d’un traitement. Prise en charge du COVID et des complications au long cours en médecine de ville et à l’hôpital; 2020; Paris.

48. Association of American Physicians and Surgeons. A guide to home-based COVID-19 treatment. 2020.

49. Min JJ-Y, Jang YJY. Macrolide therapy in respiratory viral infections. Mediators of Inflammation. 2012;2012:649570-.

50. Damle BB, Vourvahis MM, Wang EE, Leaney JJ, Corrigan BB. Clinical Pharmacology Perspectives on the Antiviral Activity of Azithromycin and Use in COVID-19. Clinical Pharmacology & Therapeutics. 2020;108(2):201–11.

51. Arshad SS, Kilgore PP, Chaudhry ZSZ, Jacobsen GG, Wang DDD, Huitsing KK, et al. Treatment with hydroxychloroquine, azithromycin, and combination in patients hospitalized with COVID-19. International journal of infectious diseases. 2020;97:396–403.

52. Cadegiani FA, Goren A, Wambier CG, McCoy J. Early COVID-19 Therapy with Azithromycin Plus Nitazoxanide, Ivermectin or Hydroxychloroquine in Outpatient Settings Significantly Reduced Symptoms Compared to Known Outcomes in Untreated Patients. medRxiv. 2020:2020.10.31.20223883.

53. Lansbury LL, Lim BB, Baskaran VV, Lim WSW. Co-infections in people with COVID-19: a systematic review and meta-analysis. The Journal of infection. 2020;81(2):266–75.

54. Nicolson GL, de Mattos GFJIJoCM. COVID-19 Coronavirus: Is Infection along with Mycoplasma or Other Bacteria Linked to Progression to a Lethal Outcome? 2020;11(05):282.

55. Lepere P EB, Yolartiran S, Escarguel C,. The Role of Macrolide Antibiotics in the Prevention of Severe COVID-19 Disease Progression Via the Disruption of Bacteria/virus Co-Operation 2020. Available from: https://papers.ssrn.com/sol3/papers.cfm?abstract_id=3712423.

56. Gehanno P. La colonisation microbienne des voies respiratoires: John Libbey Eurotext; 1995.

57. Layani Milon MP, Gras I, Valette M, Luciani J, Stagnara J, Aymard M, et al. Incidence of upper respiratory tract Mycoplasma pneumoniae infections among outpatients in RhÙne-Alpes, France, during five successive winter periods. Journal of Clinical Microbiology. 1999;37(6):1721–6.

58. Serisier DJD. Risks of population antimicrobial resistance associated with chronic macrolide use for inflammatory airway diseases. The Lancet Respiratory Medicine. 2013;1(3):262–74.

59. The World Health Organization. Antimicrobial resistance 2020 [Available from: https://www.who.int/en/news-room/fact-sheets/detail/antimicrobial-resistance.

60. Rawson TMT, Moore LSPLSP, Castro Sanchez EE, Charani EE, Davies FF, Satta GG, et al. COVID-19 and the potential long-term impact on antimicrobial resistance. Journal of antimicrobial chemotherapy. 2020;75(7):1681–4.

61. Nieuwlaat RR, Mbuagbaw LL, Mertz DD, Burrows LL, Bowdish DMEDME, Moja LL, et al. COVID-19 and Antimicrobial Resistance: Parallel and Interacting Health Emergencies. Clinical Infectious Diseases. 2020.

